# Regional variation in parental stroke history between Jeju Island and mainland Korea

**DOI:** 10.1101/2025.09.09.25335443

**Authors:** Grace Heyborne, Sujeong Kim, Choongwon Jeong, Martin Tristani-Firouzi, Edgar J. Hernandez, Melissa A. Ilardo

**Affiliations:** University of Utah; Seoul National University

**Keywords:** Familial stroke risk, cerebrovascular risk, protective factors, genetic variation

## Abstract

**Purpose:** Despite having one of the highest prevalences of hypertension in Korea, Jeju Island maintains the lowest rate of stroke mortality nationwide. This paradoxical cardiovascular health profile raises questions about the underlying factors contributing to differential stroke risk in the Jeju population.

**Methods:** We used health data from the Korea National Health and Nutrition Examination Survey (KNHANES) to investigate whether differences in regional stroke outcome can be explained by known environmental and behavioral contributors to increased cerebrovascular health risk. We applied logistic regression with backward feature selection and cross-validation to identify predictors of parental history of stroke.

**Results:** We found that individuals from Jeju Island were significantly less likely to report a parental history of stroke compared to individuals from Seoul. This finding persisted even after adjusting for known cerebrovascular risk factors, including age, sex, SBP, antihypertensive use, dietary sodium intake, BMI, and hematocrit.

**Conclusions:** Our results suggest that other hidden factors may contribute to protection against cerebrovascular disease. Given the unique population demographic history of the island, these findings prompt future analyses to explore whether the genetic variation of Jeju contributes meaningfully to stroke resilience at the population level.

## Background

Jeju Island, Korea, presents a paradox in cardiovascular health: it has one of the highest rates of hypertension in the country, yet the lowest stroke mortality rate nationwide. According to national health statistics, Jeju’s age-standardized stroke mortality rate was 24.3 per 100,000 in 2015, roughly 18% lower than the Korean national average of 29.6 per 100,000.^1^ This discrepancy was identified in a nationwide study that reported absolute differences in mortality but did not account for environmental or lifestyle contributors. As a result, the underlying causes of Jeju’s reduced stroke mortality remain unclear.

To investigate whether environmental and behavioral factors alone can explain differences in regional stroke outcome, we analyzed population-level health data from the Korea National Health and Nutrition Examination Survey (KNHANES).^2^ We compared individuals from Jeju and Seoul, the two genetically and geographically distinct Korean populations.^3^ We examined known contributors to increased cerebrovascular health risk, such as hypertension^4^, sex, age,^5^ body mass index (BMI),^6^ hematocrit^7^, alcohol consumption,^8^ smoking,^9^ and diet.^10,11^ Through this comparison, we evaluate whether the observed regional variation in stroke outcomes can be explained by known risk factors, or whether it may point to the presence of additional, unrecognized influences on population health.

## Methods

Data for this study was obtained from the Korea National Health and Nutrition Examination Survey (KNHANES)^2^, which is supported by the Korean Disease Control and Prevention Agency. The Jeju region is only reported in the years following 2017. We focused our analysis on participants from Jeju and Seoul to represent populations that have been established as genetically distinct.^3^

We used the following variables from the KNHANES dataset: age, sex, BMI, dietary sodium, dietary sugar, dietary fat, hypertension, hypertensive medication use, alcohol consumption, smoking, age SBP, DBP, hematocrit, parental history of stroke, and parental history of IHD. For details on variable definitions and PPI statement, see the Supplementary Materials.

We performed a feature selection analysis using both classical statistical modeling and regularization-based techniques. We developed six logistic regression models to evaluate and compare feature selection strategies. A ‘full model’ included all candidate predictors without prior selection. From there, we performed forward selection in which variables were added sequentially to the model based on the greatest improvement in model fit (assessed by the Akaike Information Criterion). We followed this approach with backward elimination starting from the full model, sequentially removing variables. With a final stepwise selection, we combined forward and backward procedures. Following predictor selection, we performed a LASSO regression, five-fold cross-validation, and recursive feature elimination (RFE) to identify optimal variables and remove the least informative variables. Finally, we assessed multicollinearity among candidate predictors using the Variance Inflation Factor (VIF) and further evaluated retained predictors for statistical significance (p < 0.05). For additional details on variable transformations and model development, see the Supplementary Materials.

## Results

### Regional variation in potential stroke contributors

To understand the observed regional variation in stroke mortality between Jeju and Seoul, we began by investigating which environmental variables vary regionally. We first evaluated multicollinearity using a correlation matrix (Supplementary Figure 1). We found no significant pair-wise associations and thus included all variables in subsequent analysis.

**Figure 1.**
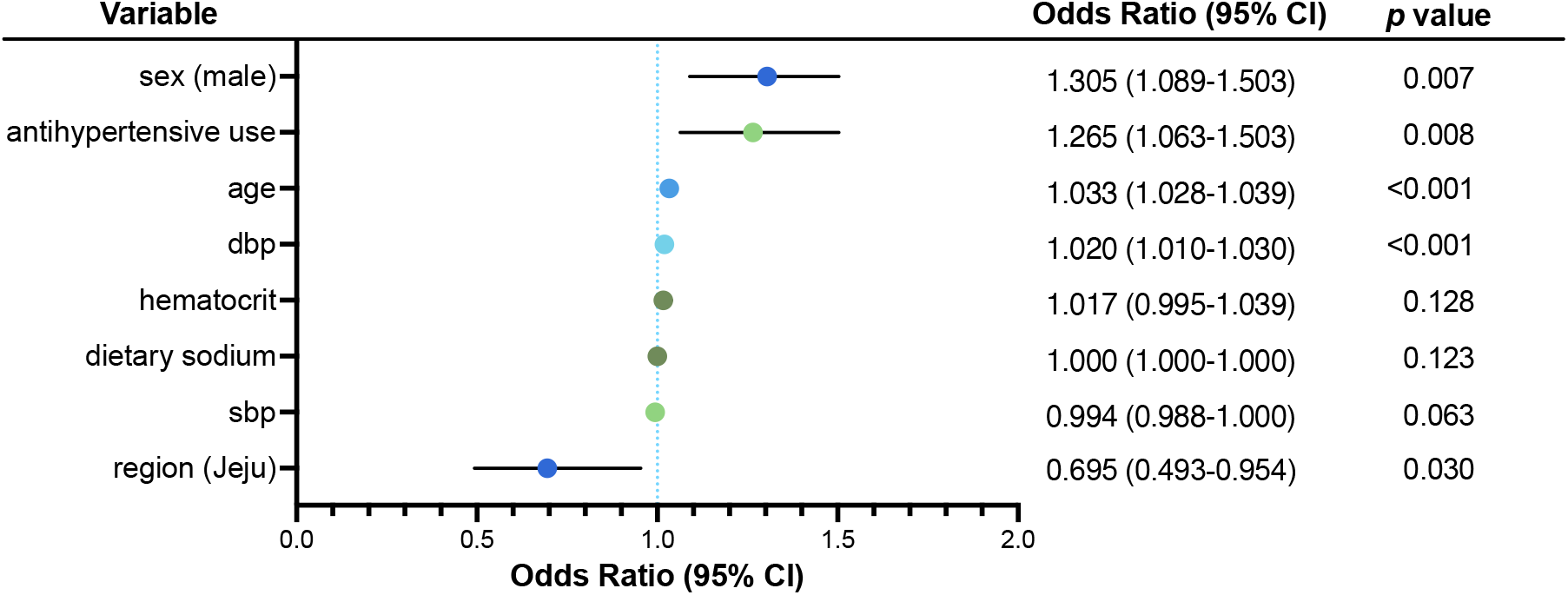
Adjusted Odds Ratios and 95% Confidence Intervals for Predictors of Parental Stroke in the Final Logistic Regression Model. All predictors were statistically significant with the exception of sodium, SBP and Hematocrit. Region as a predictor yielded an odds ratio most suggestive of a meaningful association. P-values are corrected for multiple testing using the Benjamini-Hochberg method at an FDR threshold of 0.05.

We observed multiple environmental factors that significantly differ between regions. Among the continuous traits examined, age, body mass index (BMI), systolic blood pressure (SBP), and dietary intake of fat, sodium, and sugar all differed significantly between Jeju and Seoul. Jeju participants were older (42.8 ± 23.4 y vs. 40.8 ± 21.8 y; d = 0.10) and had higher BMI (23.4 ± 4.3 vs. 22.7 ± 4.1; d = 0.18) and SBP (119.7 ± 16.8 mm Hg vs. 116.3 ± 16.7 mm Hg; d = 0.20) as well as higher hematocrit levels (42.32 ± 4.41 vs 41.92 ± 4.08; d = 0.098) than in Seoul. In contrast, Seoul participants had higher sodium intake levels (3719 ± 2544 mg vs. 3075 ± 2211 mg; d = –0.25). We found no regional differences in diastolic blood pressure, sugar, or fat (Table 1).

**Table 1.**
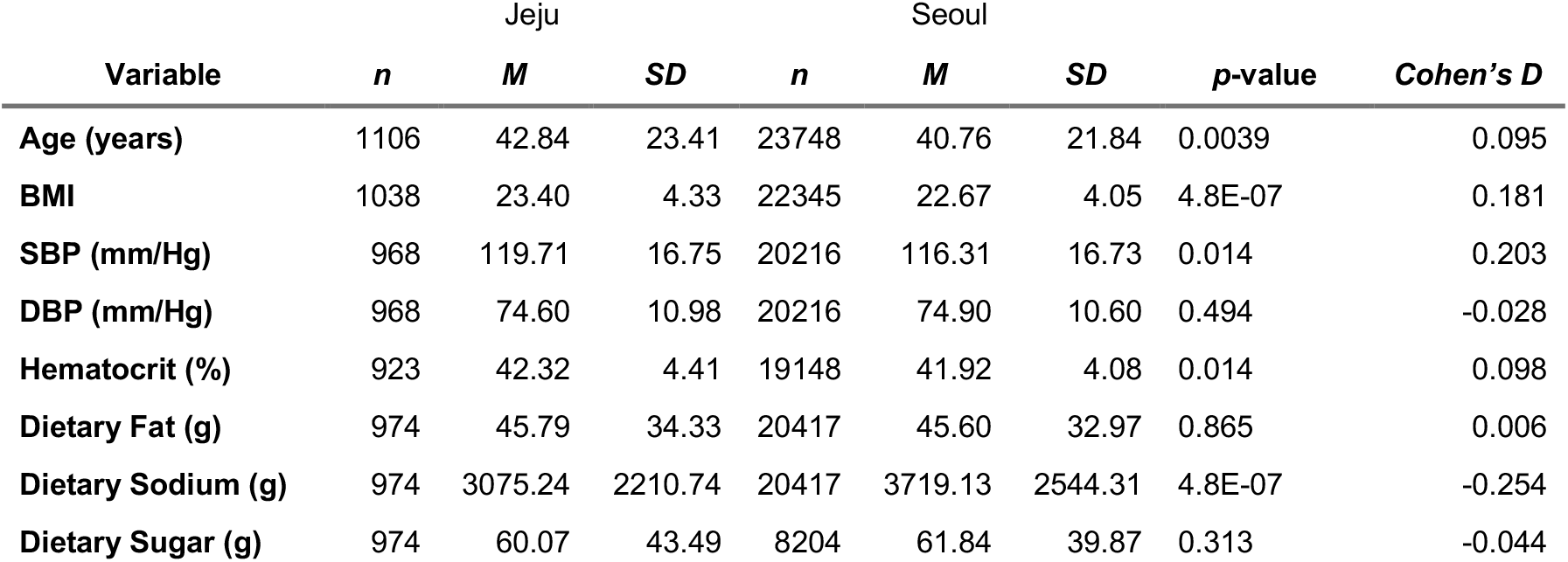
Comparison of Continuous Variables between Jeju and Seoul. Age, BMI, SBP, and dietary sodium differed significantly between Jeju and Seoul. *P*-values were corrected for multiple testing using the Benjamini-Hochberg method at a false discovery rate (FDR) threshold of 0.05.

Among discrete variables, hypertension (V = 0.037), alcohol consumption (V = 0.046), and antihypertensive medication use (V = 0.022) varied significantly by region, with higher prevalence in Jeju. These results align with previous reports^1^ that indicate a higher burden of hypertension in Jeju, likely driven by elevated SBP as DBP did not differ significantly between regions (Supplementary Figure 2). Further, despite significant regional differences in parental history of stroke (p = 0.0126; V = 0.021), there was no significant regional difference in the rate of parental ischemic heart disease (IHD), a proxy for genetic contributions to coronary artery disease (Table 2).

**Table 2.** Comparison of Discrete Variables between Jeju and Seoul. Chi-square test of association shows that parental history of stroke, hypertension, antihypertension use, and alcohol consumption differed significantly between Jeju and Seoul. *P*-values were corrected for multiple testing using the Benjamini-Hochberg method at an FDR threshold of 0.05.

| Variable | Levels | df | X<sup>2</sup> | p-value | Cramer's V |
| --- | --- | --- | --- | --- | --- |
| Sex | Male, Female | 1 | 0.72 | 0.554 | 0.006 |
| Parental Stroke | No, Yes | 1 | 6.94 | 0.014 | 0.021 |
| Hypertension (scaled) | 1,2,>3 | 2 | 25.75 | 8.96E-06 | 0.037 |
| Antihypertensive Use (scaled) | No, Yes | 1 | 11.04 | 0.00208 | 0.022 |
| Alcohol Consumption (scaled) | 1,2,3 | 2 | 37.34 | 5.44E-08 | 0.047 |
| Smoking | No, Yes | 1 | 0.01 | 0.93 | 0.001 |
| Parental IHD | No, Yes | 1 | 0.01 | 0.93 | 0.001 |

### Variable association with parental history of stroke

We next investigated which variables were associated with and/or predictive of parental history of stroke. To do this, we first evaluated multicollinearity using generalized variance inflation factors (GVIFs). All variables remained below 2.0 (GVIF threshold for elimination > 5) and were retained in the model.

We applied logistic regression with backward feature selection (AIC: 6384.4) and cross-validation to identify predictors of the binary outcome parental history of stroke. The final model retained eight predictors (Figure 1). Our results confirm that the Jeju population is associated with a 30.5% decrease in odds of a parental history of stroke (OR =0.695, 95% CI: 0.493-0.954, *p* =0.0304). Age was a significant predictor of parental history of stroke, with each additional year associated with a 3.3% increase in odds (OR = 1.03, 95% CI: 1.03–1.04, *p* < 0.001), antihypertensive use (OR = 1.26, 95% CI: 1.06–1.50, *p* = 0.0077), DBP (OR = 1.0197, 95% CI: 1.01-1.02, *p* < 0.001) and male sex (OR = 1.27, 95% CI: 1.07–1.50, *p* = 0.007) were also associated with higher odds. Although sodium, SBP, and hematocrit were retained in the model, they were not statistically significant. The intercept term was significant (OR = 0.0037, *p* < 0.001), consistent with a low baseline probability of the outcome.

## Discussion

In this study, we examined regional variation in parental history of stroke among Korean adults and found that individuals from Jeju Island were significantly less likely to report a parental history of stroke compared to individuals from Seoul. This finding persisted even after adjusting for known cerebrovascular risk factors, including age, sex, SBP, antihypertensive use, dietary sodium intake, BMI, and hematocrit. Notably, this pattern was specific to stroke; no significant regional difference was observed in reported parental history of IHD. These results align with previous epidemiological data showing lower stroke mortality in Jeju relative to Seoul,^1^ but extend these findings by demonstrating that this disparity persists even after accounting for environmental and physiological covariates.

Our results raise important questions about the potential contributors to regional differences in cerebrovascular health. While Seoul participants exhibited higher sodium intake, an established risk factors for vascular disease, dietary sodium was not found to be a significant predictor of parental stroke history in our models. Although hypertension was more prevalent in Jeju, this difference was driven largely by elevated SBP, with no significant regional difference in DBP. However, only DBP was found to be a predictor of parental history of stroke in our model. This distinction is particularly notable given that previous evolutionary genomic analyses in the Jeju population identified variants under positive selection associated with attenuated increases in DBP during breath-hold diving.^3^

Jeju Island represents a genetically and culturally distinct population within Korea.^12^ The island’s population has been shaped by its demographic history, including long periods of relative isolation and admixture events such as Mongolian influx during the 13th century. The genetics of Jeju may also have been influenced by a unique cultural practice: generations of women who engage in extreme breath-hold diving.^12-14^ This practice, which often continues through pregnancy, may have imposed selective pressures on cardiovascular physiology. Indeed, a recent study identified a genetic variant under selection in Jeju Islanders that was associated with a ∼10 mmHg reduction in DBP per allele during diving.^3^ This variant functions as an expression quantitative trait locus (eQTL) for FcγRIIA, an IgG2 receptor with a known role in modulating vascular inflammation.^15^ FcγRIIA may reduce diastolic hypertension by dampening inflammatory responses, thus potentially buffering against the hypertensive effects of the dive reflex. Although the direct relationship between this variant and stroke risk remains unclear, its known influence on DBP provides a plausible biological pathway through which genetic adaptation could confer cerebrovascular protection in Jeju Islanders.

These findings suggest that Jeju Islanders may possess protective mechanisms against cerebrovascular disease that are not captured by conventional risk factors. The observation that stroke family history is lower in Jeju despite higher hypertension prevalence highlights a potential disconnect between blood pressure levels and downstream disease risk and supports the hypothesis that distinct physiological or genetic buffering mechanisms may be at play. Future studies should explore whether the genetic variants under selection in Jeju contribute meaningfully to stroke resilience at the population level. Functional characterization of the FcγRIIA eQTL, as well as its interaction with inflammatory and vascular pathways, will also be essential for determining its clinical relevance.

These findings should be interpreted in light of several limitations. Lifestyle and environmental factors not captured by the KNHANES survey, such as healthcare access, rural versus urban living, and patterns of physical activity, may influence stroke risk and detection. Additionally, the KNHANES dataset reflects a population imbalance, with only 1,106 Jeju individuals compared to 23,748 from Seoul between 2017 and 2022, potentially impacting statistical power. Future studies should aim to address these limitations by incorporating more balanced sampling and finer-grained environmental and behavioral data.

## Data Availability

All data produced in the present study are available upon reasonable request to the authors.

## Data and Code Availability

All original code will deposited to github and will be made publicly available as of the date of publication.

