## Supplementary Materials for "Regional variation in parental stroke history between Jeju Island and mainland Korea"

**Supplementary Figures**

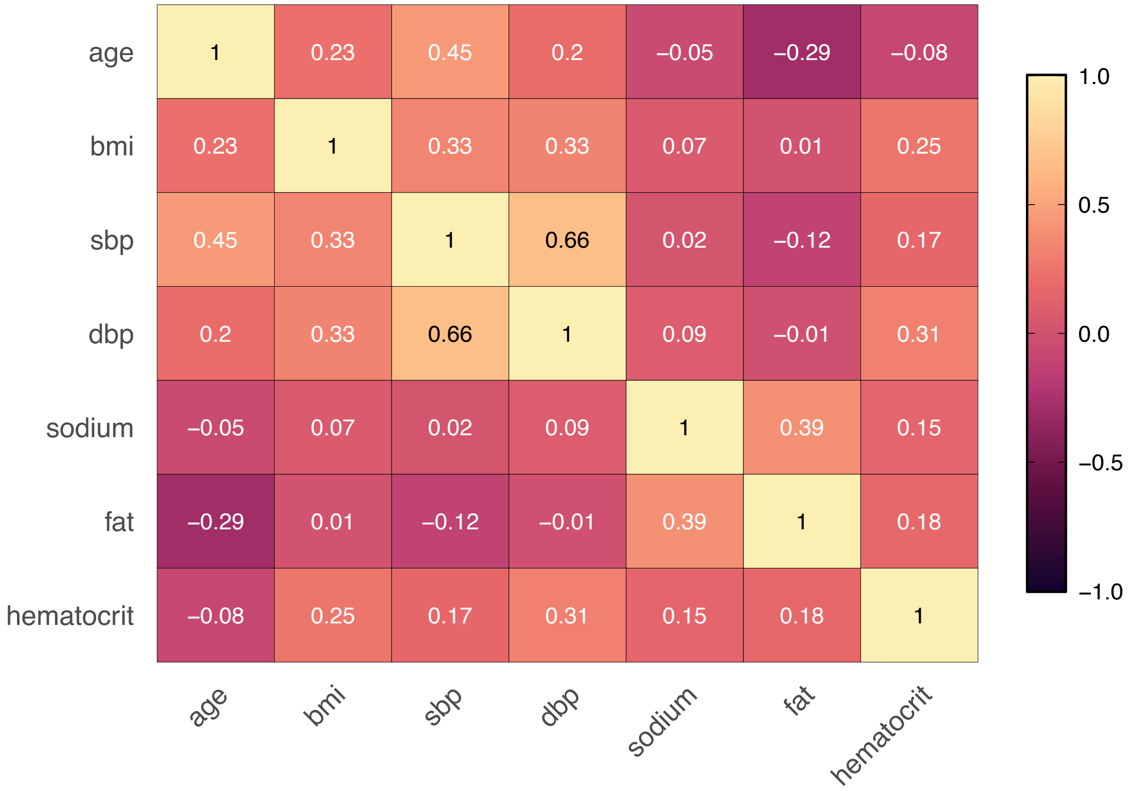


**Supplementary Figure 1**. Correlation Matrix Heatmap Between Potentially Collinear Variables (calculated by Pearson R). No significant pair-wise associations were found and therefore all variables were included in the analysis.

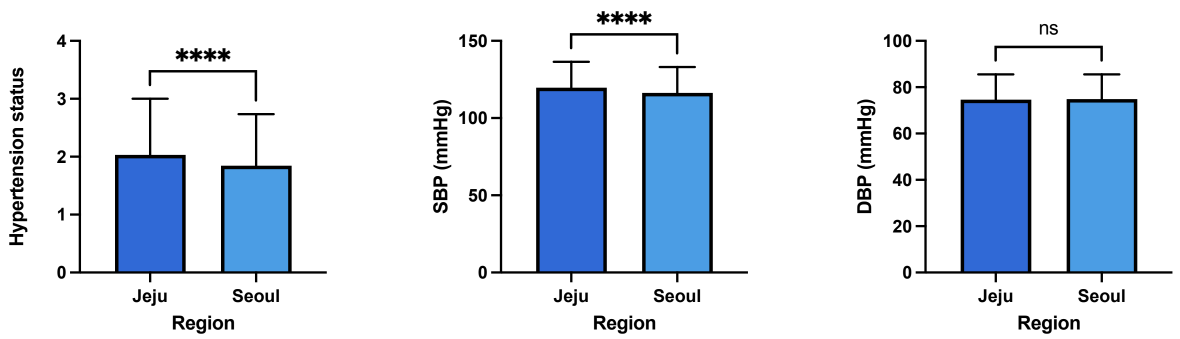


**Supplementary Figure 2**. Hypertension, SBP, and DBP by Region. The average hypertension status differs significantly between Jeju and Seoul. SBP differs significantly between regions while DBP does not, suggesting that difference in hypertension rate is driven by SBP rather than DBP. **** $p < 0.0001$.

**Supplementary Methods**

Detailed Description of Variables

Smoking, BMI, dietary composition, hematocrit, and SBP/DBP are measured on a continuous scale. Smoking quantity denotes the average number of cigarettes per day, and BMI is calculated by dividing reported weight by reported height. Hematocrit is denoted as the percentage of red blood cells per sample. Dietary composition for fat, sodium, and sugar was obtained from survey responses that detail diet composition within the prior 24 hours. For both SBP and DBP, blood pressure was recorded three separate, consecutive times before averaging the 2$^{nd}$ and 3$^{rd}$ readings to obtain a 'final' blood pressure reading.

The remaining variables—antihypertensive use, alcohol consumption, parental stroke, parental ischemic heart disease, and hypertension status are all ranked variables. Antihypertensive medication use, smoking, parental IHD, and parental stroke are coded as binary yes/no variables. Alcohol consumption is ranked on a 1-3 scale. A response of 1 indicates no alcohol consumption at all in the past year, 2 indicates alcohol consumption >1-4 times a month, and 3 indicates 2-4 times a week. Lastly, hypertension status ranges from 1-3, where 1 indicates normal blood pressure, 2 indicates prehypertension (elevated SBP between 120-129, but normal DBP of less than 80), 3 indicates either hypertension pre-stage and hypertension (pre-stage hypertension: SBP between 130-139 and DBP of 80-89, hypertension: SBP at or above 140 and DBP at or above 90).

Detailed Description of Statistical Analyses

Variable transformations: Categorical variables (e.g., sex, region, use of antihypertensive medications, smoking and drinking status) were transformed into indicator variables using one-hot encoding. This allowed for seamless integration into penalized regression models while preserving the interpretability of binary covariates. The outcome variable was parent stroke was coded as a binary 0/1 (no/yes).

Lasso regression and five-fold cross-validation: This regression penalized logistic regression model with an L1 penalty which was fitted using the glmnet package. Five-fold cross-validation was performed to identify the optimal regularization parameter ($\lambda$), and variables with non-zero coefficients at the optimal $\lambda$ were retained. To avoid double standardization, predictors were manually scaled, and the standardize = FALSE option was set in the modeling function. Lastly, an Elastic Net Regression was performed. This regression is similar to LASSO but incorporates both L1 and L2 penalties ($\alpha = 0.5$). Five-fold cross-validation was used to select the optimal $\lambda$ value, and manually scaled predictors were used.

REA: Recursive feature elimination (RFE) was conducted using random forest as the base classifier via the caret package in R. The process recursively evaluated subsets of predictors by iteratively removing the least informative variables based on coefficient magnitude. Model performance was assessed through 5-fold cross-validation, and the optimal subset was selected based on minimal cross-validated error.

VIF: Prior to final model fitting, multicollinearity among candidate predictors was assessed using the Variance Inflation Factor (VIF). Predictors with a VIF exceeding a threshold of 10 were flagged as potentially collinear. These variables were either removed or carefully monitored in subsequent model fitting. VIF assessment was conducted on the full model using the car package in R. Each model was evaluated using AIC and the Bayesian Information Criterion (BIC) to assess model fit.

PPI Statement

The Korean public contributed to this study through their participation in the Korea National Health and Nutrition Examination Survey (KNHANES), an annual survey designed to provide accurate and timely nationwide health statistics. Prior Korean public health reports have documented regional differences in the prevalence of both hypertension and stroke, conditions that pose critical challenges to healthcare systems. Motivated by these findings, this study aimed to build upon existing health data by examining environmental and behavioral contributors to cerebrovascular risk. Parental stroke history was selected as an outcome measure to access regional variation in stroke burden. While members of the public were not involved in the design of this study directly, its objectives align with public health interests and aim to inform population-level health disparities.
